# Avoiding being the “busy fool”: How general practitioners perceive and engage with a prescribing safety and quality dashboard

**DOI:** 10.64898/2026.05.07.26352633

**Authors:** Anais Essilini, Barbara Clyne, Tom Fahey, Frank Moriarty, Michelle Flood, Claire Gorry, Caroline McCarthy

## Abstract

**Background:** Interactive dashboards can support safe prescribing but effectiveness depends on user engagement. The research team developed a prescribing safety dashboard, deployed in 27 Irish general practices. Trend graphs tracked prescribing changes (2019-2025) by practices across key metrics. This study explored how GPs engaged with the dashboard and their perceptions of using routine data for prescribing feedback.

**Methods:** Prescribers from participating practices were invited to online interviews (May-August 2025). A think-aloud exercise involved participants verbalising their thoughts while navigating the dashboards, followed by a semi-structured interview exploring views on safe prescribing, feedback and data access. Interviews were recorded, auto-transcribed and manually reviewed for accuracy. Think-aloud data were analysed deductively using a sense-making framework, interviews analysed inductively, and findings triangulated to refine themes.

**Results:** Nine general practitioners (GPs) from eight practices participated. Themes were organised into four categories: (1) Perceptions of open data, (2) Perceptions of feedback, (3) Dashboard engagement, and (4) High-quality prescribing. Most were in favour of open data and transparency but some feared misuse. GPs valued feedback but reported workload as a barrier. Engagement with the dashboard was mainly interpretative, focused on data meaning in the context of their practice. GPs showed a strong emotional dimension to engagement and also described intended actions in response to what they saw. Finally, high-quality prescribing was mainly viewed as avoiding harm.

**Conclusions:** GPs valued and engaged with dashboard feedback but workload competed with time for reflection and action-highlighting the need for practical, streamlined tools and nudges to support engagement.

**Key messages:**

- Audit and feedback, such as that delivered through interactive dashboards has a small but significant effect on professional behaviours such as prescribing, but user engagement influences effectiveness.
- Irish GPs engaged with a prescribing safety and quality dashboard in a reflective and contextual way and garnered rich insights on their prescribing.
- GPs valued feedback and showed a strong emotional attachment to their performance, but felt workload competed with time for reflection and action.
- With advances in data infrastructure, it is possible to provide interactive prescribing feedback in real time. However, the way feedback is designed and delivered plays a crucial role in supporting engagement. Dashboards and related behavioural interventions should be co-designed with prescribers to maximise engagement.

## Introduction

Medication-related harm is an important contributor to morbidity and mortality worldwide. Approximately 5% of all hospital admissions are due to preventable drug-related morbidity (1), rising to 9% in older adults (2). The World Health Organization’s Third Global Patient Safety Challenge, *Medication Without Harm*, aims to reduce severe, avoidable medication-related harm (3). In addition, there is growing recognition of the phenomenon of overprescribing and its social and economic consequences, even when patients do not experience direct medication-related harm (4, 5).

These challenges are particularly relevant to primary care, where most prescribing occurs. Well-developed primary care systems are recognised as essential for the equitable distribution of care, including the prescription of medicines to treat and prevent disease (6). This includes the acute prescribing for intercurrent illness, and the initiation and continuation of long-term therapies, that may have been started in secondary care. Prescribing in primary care typically occurs under time pressure and in relative professional isolation, with limited opportunities for reflection or peer discussion and in the context of a fragmented and single-disease orientated healthcare system (7). Evidence suggests that general practitioners (GPs) value feedback to support safe, appropriate prescribing (8).

Audit and feedback has a small effect on professional behaviours such as prescribing (9), and the degree of engagement with the feedback is a key determinant of the effectiveness of these interventions (10). Future work should focus on systematically developing and testing feedback interventions with attention to their design and delivery so that are tailored to the needs of end users (10). Advances in primary care data infrastructure have made it increasingly feasible to deliver ongoing, comparative feedback on prescribing quality. A prominent example is *OpenPrescribing.net*, which provides regularly updated data on prescribing patterns for general practices in NHS England (11). There is emerging evidence that interactive prescribing dashboards such as these can help support high-quality prescribing in general practice (12). The aim of this study was thus to explore how GPs used a prescribing safety and quality dashboard developed for Irish general practice. The first objective was to explore how prescribers interacted with and interpreted the dashboards. The second was to examine perceptions of using routine data as feedback to support safe prescribing, including its usefulness and acceptability.

## Methods

This qualitative study is reported in line with COREQ guidelines (Appendix 1). The prescribing indicators used in the dashboard were informed by a prior pilot study (13), and the dashboard development and study protocol are described elsewhere (14).

### Setting

This study was conducted in the Republic of Ireland, where approximately 30% of the population are eligible for the General Medical Services (GMS) scheme, which entitles individuals to free GP visits and medicines (with a €1.50 prescription charge per medicine up to a maximum of €15 per month) (15). GP prescribing for GMS patients is captured in the Health Service Executive Primary Care Reimbursement Service (PCRS) database, whereas prescribing outside these schemes is not routinely recorded. Participating practices currently receive limited prescribing feedback, typically in the form of quarterly paper reports of GMS scheme antibiotic and benzodiazepine prescribing.

### Dashboard description

The dashboard was developed between October 2024 and January 2025 by the senior author (CMC) in collaboration with MedVault a private data analytics company that supports practices in using their data to manage reimbursement claims. The dashboard comprised seven tabs; prescribing of HSE Preferred Drugs (16), opioids, benzodiazepines, top 90% drugs (DU90%), high-risk non-steroidal anti-inflammatory drugs (NSAIDs), high-dose proton pump inhibitors (PPIs) and antibiotics. All tabs, apart from the ‘Top 90% prescribed drugs’, displayed comparative trend graphs. These allowed users to track annual prescribing changes from 2019 to 2025 and to compare their rates with other practices. The ‘Top 90% prescribed drugs’ tab presented a bar chart showing, in decreasing frequency, the prescribing volume of each drug within the DU90% segment for that practice, Figure 1.

**Figure 1.**
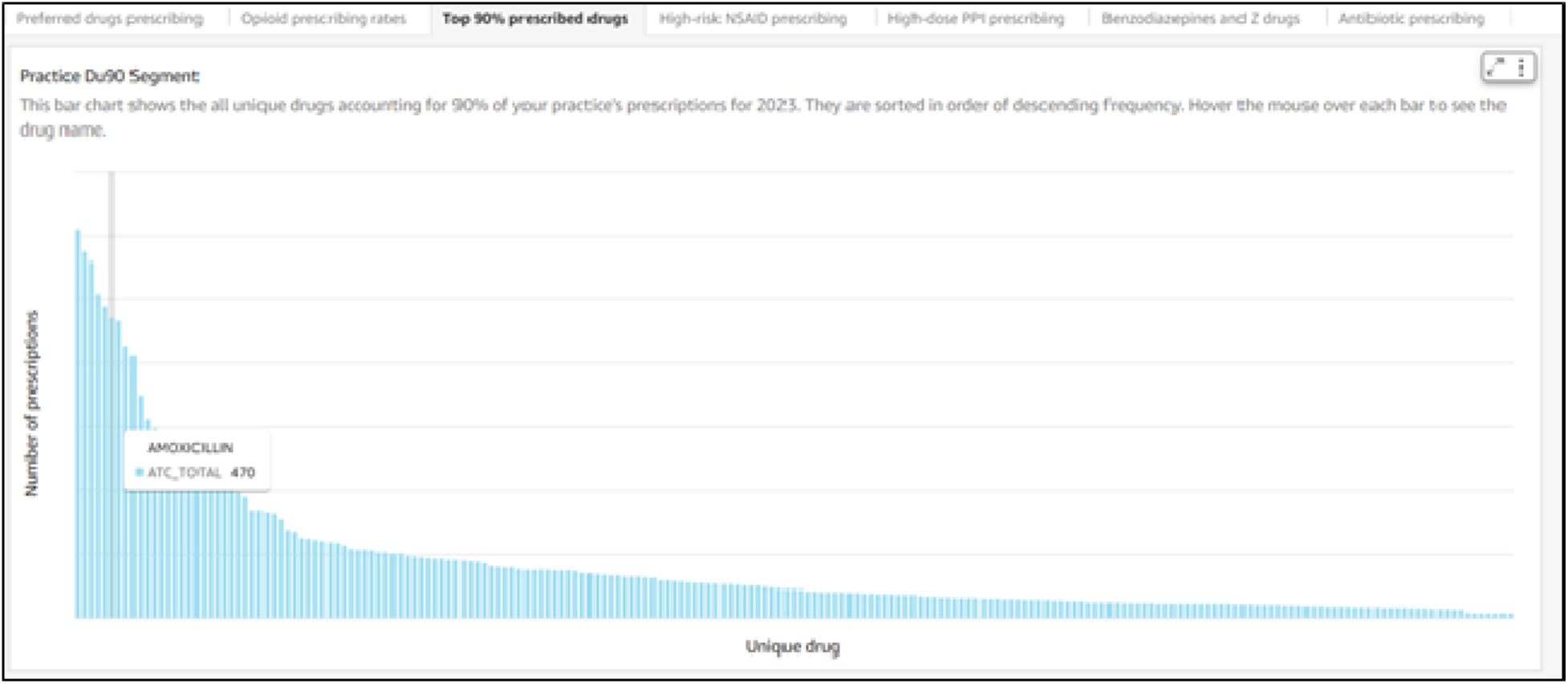
Practice DU90%

### Study population

Twenty-seven of 153 eligible MedVault practices signed up to receive the dashboards. The median number of core prescribers (defined as those responsible for ≥5% of all prescriptions in a given clinic–year) per practice was 5 (IQR 4–6), indicating included practices were slightly larger than national averages (17). The median GMS-to-private prescription ratio was 0.33 (IQR 0.30–0.37), closely reflecting the national proportion of GMS eligibility (18). Twenty-two of 27 practices consented for further contact and were invited to participate in this qualitative study by email and follow-up telephone call.

### Data Collection

Interviews were conducted by the senior author, an Irish female GP and health services researcher (CMC) on Microsoft Teams where the participant screen-shared while they accessed the dashboards. The interview began with the think-aloud exercise which involved four predefined tasks exploring, 1) antibiotic prescribing, 2) high-risk NSAID use, 3) DU90% and 4) an open-ended task where participants could explore any remaining sections of the dashboard. After the third interview, a fifth task-focused on preferred drug prescribing-was added to the protocol as participants engaged with it in varied and reflective ways for the open-ended task, prompting its formal inclusion. The second part of the process was a semi-structured interview where views around safe prescribing (https://zenodo.org/records/15632494), feedback on prescribing and access to data were explored. Interviews were recorded, auto-transcribed and manually reviewed for accuracy. Third-party references and identifying details were redacted. All participants were given the opportunity to review transcripts for accuracy and intended meaning.

### Analysis

Coding for the think-aloud process was conducted by the senior author (CMC) and for the semi-structured interviews by the first author, a French female health services researcher (AE). During the think-aloud coding the screencast video was viewed alongside the transcript and the transcript annotated with relevant details (for example describing what the GP was looking at or clicking on). A deductive coding framework based on Nielsen’s five quality components of usability was originally planned, to analyse the think-aloud exercise (19). However, a sense-making approach was ultimately adopted as this better captured the interpretative and cognitive work involved and had been adapted for a similar analysis which explored teacher interactions with visual analytics dashboards (20, 21). Annotated transcripts were analysed deductively using three predefined categories; emotional, interpretive and intentional engagement-representing affective responses to data, efforts to explain or contextualise it, and indications of intended action, respectively. Analysis for the semi-structured interviews was inductive, with codes generated from the data. Both analyses were conducted using QSR International’s NVivo version 15.0.0 for the think-aloud and version 1.6.1 for the semi-structured interviews. The independently coded think-aloud and interview data were subsequently compared and discussed, allowing triangulation of insights and refinement of overarching themes.

## Results

Nine GPs were recruited from eight practices. At interview one GP could not access the dashboard and so think-aloud data was collected from eight GPs and semi-structured interview data from nine. Recruited GPs were predominately female (7/9) and practice partners (6/9). Interviews lasted on average 40 minutes (25-54 minutes). Themes were organised into four overarching categories: (1) Perceptions of open data, (2) Perceptions of feedback, (3) Dashboard engagement, and (4) Safe and high-quality prescribing. Figure 2 illustrates the relationships between these categories, showing how perceptions of data and feedback influence engagement with the dashboard and, ultimately, approaches to safe, high-quality prescribing. Illustrative quotes are presented in Table 1 and are referred to throughout by their quote number (e.g., Q1).

**Figure 2.**
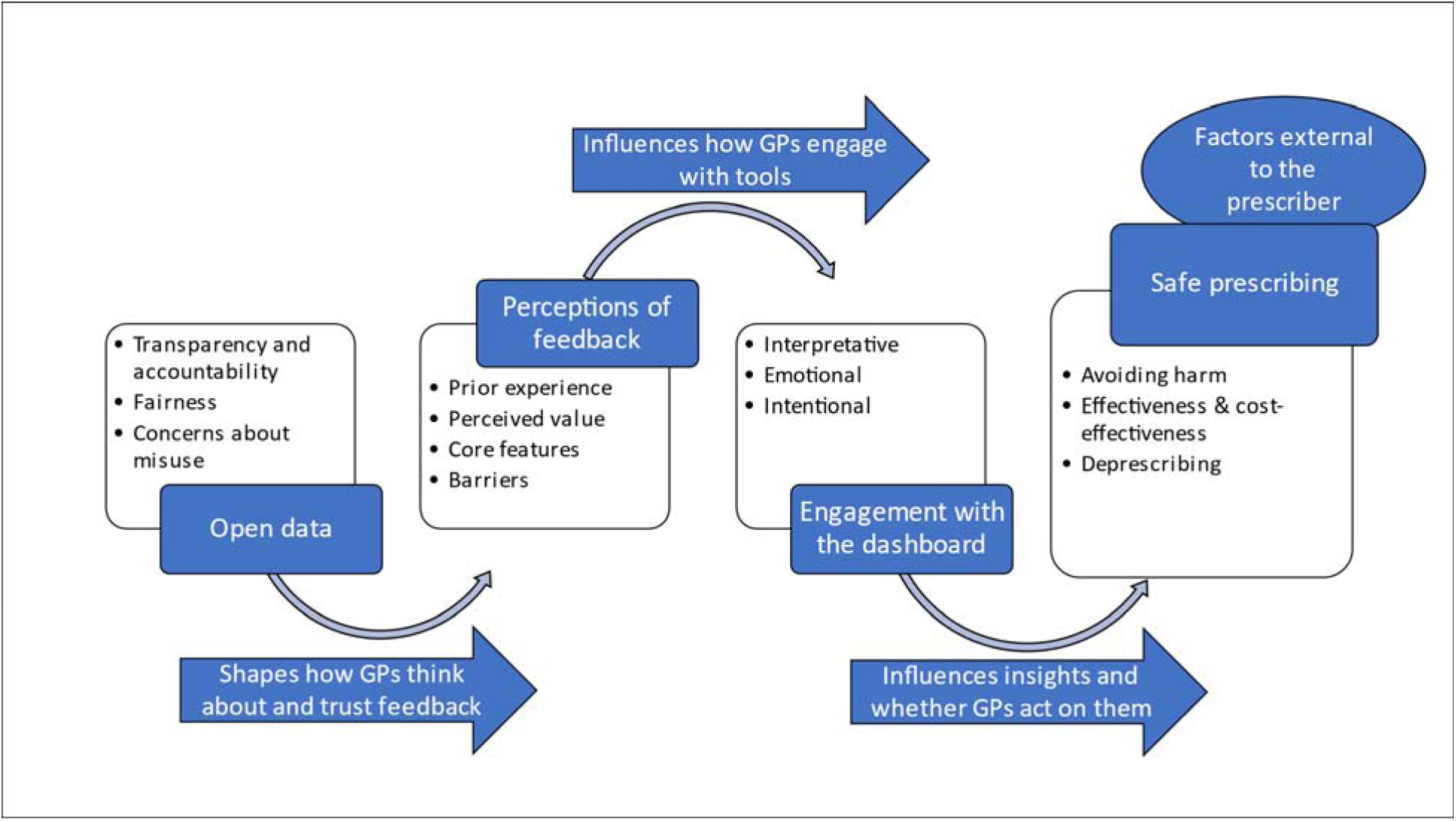
Thematic map describing factors shaping engagement with prescribing dashboards

**Table 1.**
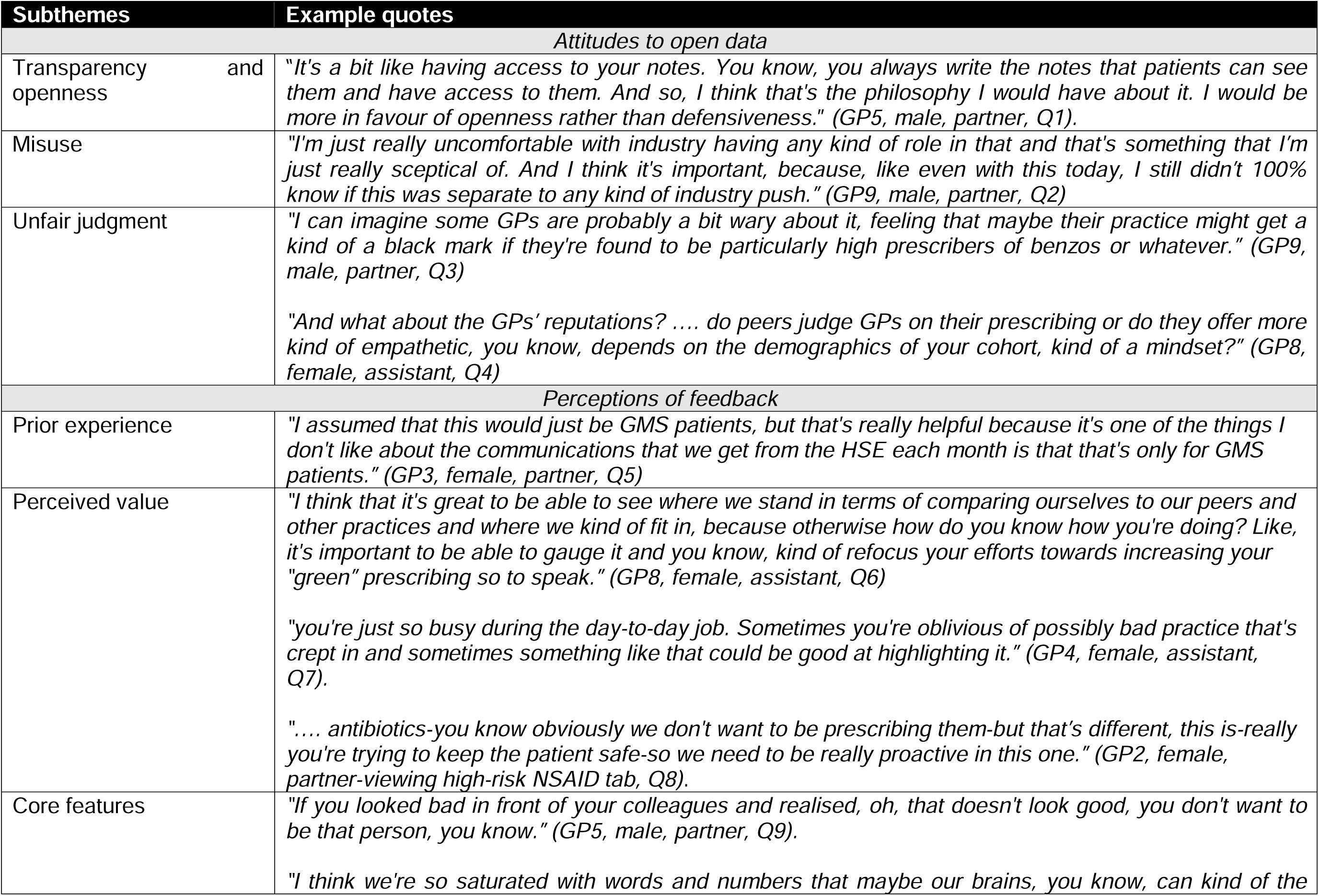
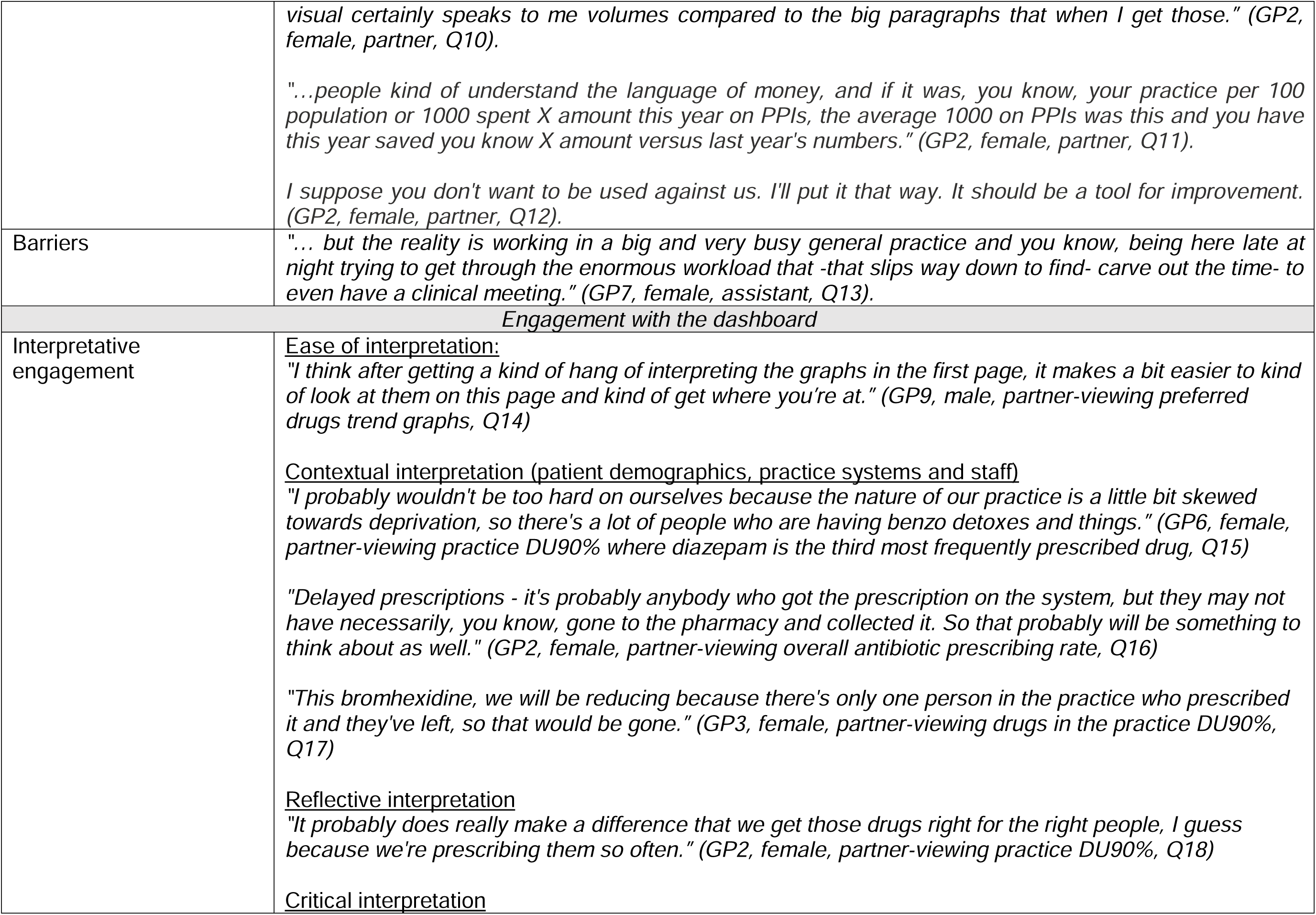
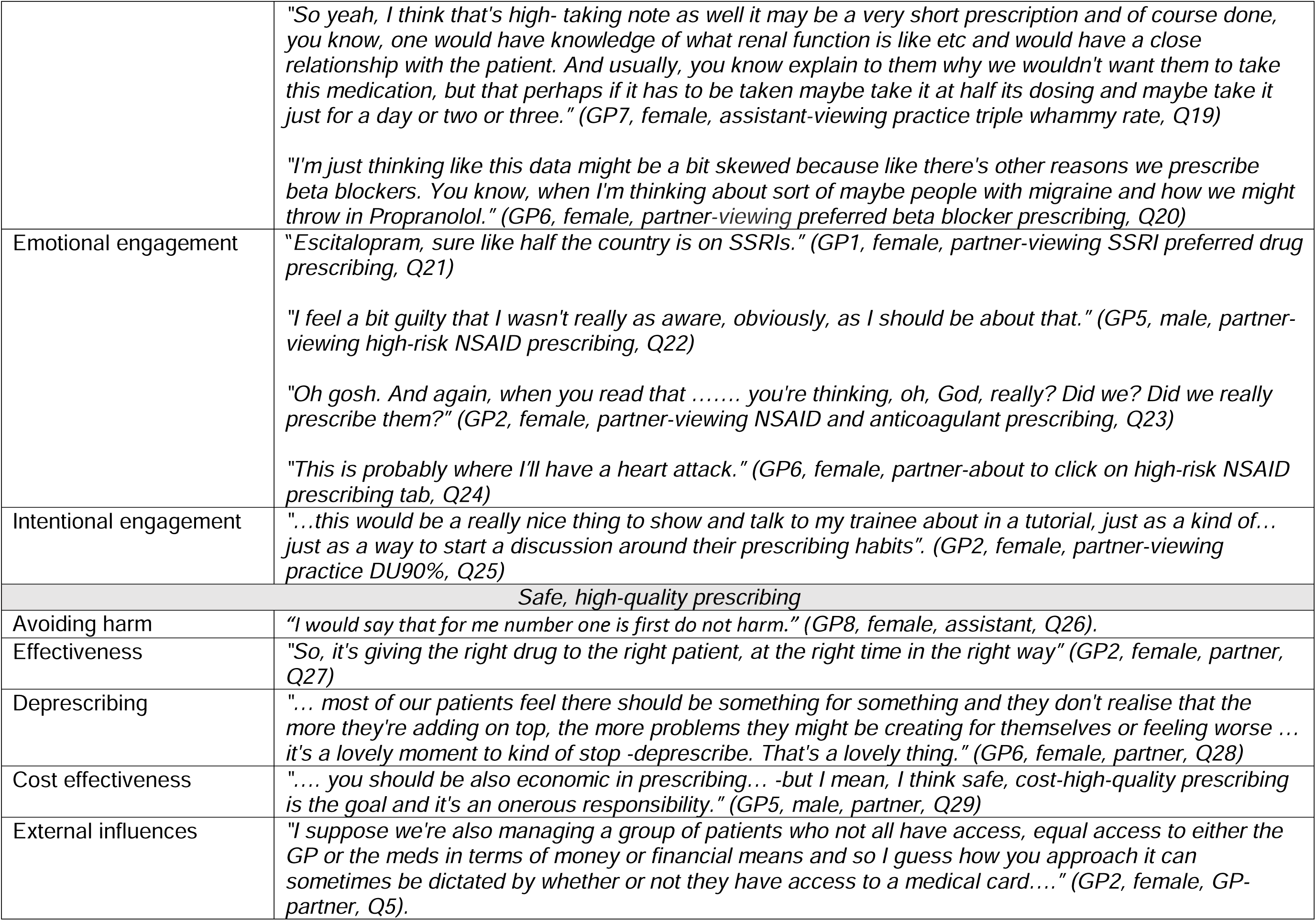
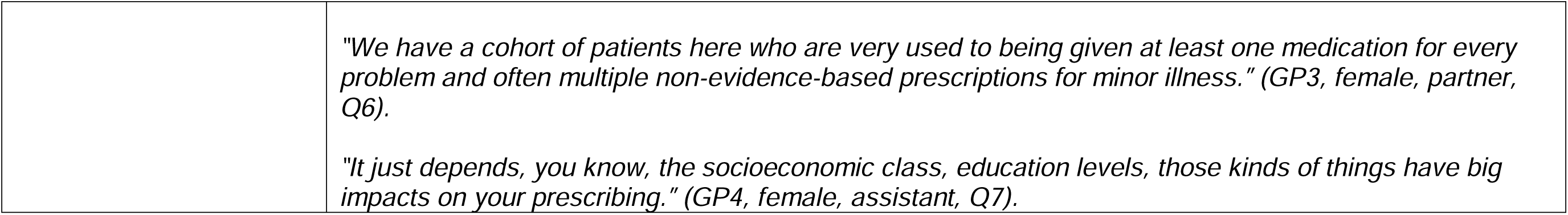
Categories, sub-themes, and supporting quotes.

| **Subthemes** |  | **Example quotes** |
| --- | --- | --- |
| *Attitudes to open data* |  |  |
| Transparency and openness |  | *"It's a bit like having access to your notes. You know, you always write the notes that patients can see them and have access to them. And so, I think that's the philosophy I would have about it. I would be more in favour of openness rather than defensiveness." (GP5, male, partner, Q1).* |
| Misuse |  | *"I'm just really uncomfortable with industry having any kind of role in that and that's something that I'm just really sceptical of. And I think it's important, because, like even with this today, I still didn't 100% know if this was separate to any kind of industry push." (GP9, male, partner, Q2)* |
| Unfair judgment |  | *"I can imagine some GPs are probably a bit wary about it, feeling that maybe their practice might get a kind of a black mark if they're found to be particularly high prescribers of benzos or whatever." (GP9, male, partner, Q3)*<br>*"And what about the GPs' reputations? .... do peers judge GPs on their prescribing or do they offer more kind of empathetic, you know, depends on the demographics of your cohort, kind of a mindset?" (GP8, female, assistant, Q4)* |
| *Perceptions of feedback* |  |  |
| Prior experience |  | *"I assumed that this would just be GMS patients, but that's really helpful because it's one of the things I don't like about the communications that we get from the HSE each month is that that's only for GMS patients." (GP3, female, partner, Q5)* |
| Perceived value |  | *"I think that it's great to be able to see where we stand in terms of comparing ourselves to our peers and other practices and where we kind of fit in, because otherwise how do you know how you're doing? Like, it's important to be able to gauge it and you know, kind of refocus your efforts towards increasing your "green" prescribing so to speak." (GP8, female, assistant, Q6)*<br>*"you're just so busy during the day-to-day job. Sometimes you're oblivious of possibly bad practice that's crept in and sometimes something like that could be good at highlighting it." (GP4, female, assistant, Q7).*<br>*".... antibiotics-you know obviously we don't want to be prescribing them-but that's different, this is-really you're trying to keep the patient safe-so we need to be really proactive in this one." (GP2, female, partner-viewing high-risk NSAID tab, Q8).* |
| Core features |  | *"If you looked bad in front of your colleagues and realised, oh, that doesn't look good, you don't want to be that person, you know." (GP5, male, partner, Q9).*<br>*"I think we're so saturated with words and numbers that maybe our brains, you know, can kind of the* |

|  |  |
| --- | --- |
|  | *visual certainly speaks to me volumes compared to the big paragraphs that when I get those.” (GP2, female, partner, Q10).*<br>*“...people kind of understand the language of money, and if it was, you know, your practice per 100 population or 1000 spent X amount this year on PPIs, the average 1000 on PPIs was this and you have this year saved you know X amount versus last year's numbers.” (GP2, female, partner, Q11).*<br>*I suppose you don't want to be used against us. I'll put it that way. It should be a tool for improvement. (GP2, female, partner, Q12).* |
| Barriers | *“... but the reality is working in a big and very busy general practice and you know, being here late at night trying to get through the enormous workload that -that slips way down to find- carve out the time- to even have a clinical meeting.” (GP7, female, assistant, Q13).* |
| **Engagement with the dashboard** |  |
| Interpretative engagement | <u>Ease of interpretation:</u><br>*“I think after getting a kind of hang of interpreting the graphs in the first page, it makes a bit easier to kind of look at them on this page and kind of get where you're at.” (GP9, male, partner-viewing preferred drugs trend graphs, Q14)*<br><u>Contextual interpretation (patient demographics, practice systems and staff)</u><br>*“I probably wouldn't be too hard on ourselves because the nature of our practice is a little bit skewed towards deprivation, so there's a lot of people who are having benzo detoxes and things.” (GP6, female, partner-viewing practice DU90% where diazepam is the third most frequently prescribed drug, Q15)*<br>*“Delayed prescriptions - it's probably anybody who got the prescription on the system, but they may not have necessarily, you know, gone to the pharmacy and collected it. So that probably will be something to think about as well.” (GP2, female, partner-viewing overall antibiotic prescribing rate, Q16)*<br>*“This bromhexidine, we will be reducing because there's only one person in the practice who prescribed it and they've left, so that would be gone.” (GP3, female, partner-viewing drugs in the practice DU90%, Q17)*<br><u>Reflective interpretation</u><br>*“It probably does really make a difference that we get those drugs right for the right people, I guess because we're prescribing them so often.” (GP2, female, partner-viewing practice DU90%, Q18)*<br><u>Critical interpretation</u> |
|  | *"So yeah, I think that's high- taking note as well it may be a very short prescription and of course done, you know, one would have knowledge of what renal function is like etc and would have a close relationship with the patient. And usually, you know explain to them why we wouldn't want them to take this medication, but that perhaps if it has to be taken maybe take it at half its dosing and maybe take it just for a day or two or three." (GP7, female, assistant-viewing practice triple whammy rate, Q19)*<br>*"I'm just thinking like this data might be a bit skewed because like there's other reasons we prescribe beta blockers. You know, when I'm thinking about sort of maybe people with migraine and how we might throw in Propranolol." (GP6, female, partner-viewing preferred beta blocker prescribing, Q20)* |
| Emotional engagement | *"Escitalopram, sure like half the country is on SSRIs." (GP1, female, partner-viewing SSRI preferred drug prescribing, Q21)*<br>*"I feel a bit guilty that I wasn't really as aware, obviously, as I should be about that." (GP5, male, partner-viewing high-risk NSAID prescribing, Q22)*<br>*"Oh gosh. And again, when you read that ..... you're thinking, oh, God, really? Did we? Did we really prescribe them?" (GP2, female, partner-viewing NSAID and anticoagulant prescribing, Q23)*<br>*"This is probably where I'll have a heart attack." (GP6, female, partner-about to click on high-risk NSAID prescribing tab, Q24)* |
| Intentional engagement | *"...this would be a really nice thing to show and talk to my trainee about in a tutorial, just as a kind of... just as a way to start a discussion around their prescribing habits". (GP2, female, partner-viewing practice DU90%, Q25)* |
| *Safe, high-quality prescribing* |  |
| Avoiding harm | *"I would say that for me number one is first do not harm." (GP8, female, assistant, Q26).* |
| Effectiveness | *"So, it's giving the right drug to the right patient, at the right time in the right way" (GP2, female, partner, Q27)* |
| Deprescribing | *"... most of our patients feel there should be something for something and they don't realise that the more they're adding on top, the more problems they might be creating for themselves or feeling worse ... it's a lovely moment to kind of stop -deprescribe. That's a lovely thing." (GP6, female, partner, Q28)* |
| Cost effectiveness | *".... you should be also economic in prescribing... -but I mean, I think safe, cost-high-quality prescribing is the goal and it's an onerous responsibility." (GP5, male, partner, Q29)* |
| External influences | *"I suppose we're also managing a group of patients who not all have access, equal access to either the GP or the meds in terms of money or financial means and so I guess how you approach it can sometimes be dictated by whether or not they have access to a medical card...." (GP2, female, GP-partner, Q5).* |
|  | *"We have a cohort of patients here who are very used to being given at least one medication for every problem and often multiple non-evidence-based prescriptions for minor illness." (GP3, female, partner, Q6).*<br>*"It just depends, you know, the socioeconomic class, education levels, those kinds of things have big impacts on your prescribing." (GP4, female, assistant, Q7).* |

### Attitudes to open prescribing data

GPs held conflicting views on open prescribing data. Most had not given much thought to the issue prior to the interview. Overall, GPs were in favour of transparency *(GP5, male, partner, Q1)*. However, some expressed concern about potential misuse by third parties, such as patients seeking practices with higher opioid prescribing, or pharmaceutical companies using data for marketing purposes *(GP9, male, partner, Q2)*. Some GPs expressed concern about being unfairly judged- particularly for benzodiazepines- by those unfamiliar with the context of their practice. Others were less concerned with external judgement and felt that patients’ views mattered most, as they are the ones taking the medicines, *“… it kind of doesn’t really matter what we think as doctors, but we need to keep the patient as the focus.” (GP3, female, partner)*. With respect to data protection, that same GP described what they saw as an oxymoron; efforts to “protect” data can, in practice, prevent meaningful scrutiny and public benefit: *“It’s like, really, who are we protecting?. The easy thing to do is to say*-*“that will be identifiable”* -*but if we’ve removed identifiers and we’re looking at raw data, I don’t see what the issue is. I expect the powers that be don’t want to see some of the data because it’s so concerning.”*

### Perceptions of feedback

GPs valued receiving feedback but reported limited experience of it. All GPs felt comparative benchmarking was an important component and some GPs had specific suggestions for other aspects that should be included, particularly metrics that display cost. Workload and competing, more pressing demands were the most cited barriers.

### Prior experience

Most GPs were aware of and welcomed the quarterly paper-based reports on benzodiazepine and antibiotic prescribing from the PCRS, which cover only GMS patients, however GP assistants who did not have GMS contracts had little experience of these reports. Most GPs believed they did not get sufficient prescribing feedback and would welcome more. When discussing feedback, several GPs focused on the immediate, interpersonal feedback they receive from community pharmacists, *“…the biggest safety net for us is the pharmacy itself…….they’re kind of the gatekeepers because if things slip through the net accidentally, they’re always the first to flag it.” (GP4, female, assistant)*.

### Perceived value

Acknowledging heavy workloads, GPs felt feedback offered an opportunity to address sub-optimal practice that may have crept in and avoid becoming a *“busy fool.” (GP1, female, partner).* Overall GPs viewed this dashboard positively, particularly that it included data for all prescribing, not just the GMS scheme. Some GPs acknowledged that different metrics may carry more importance. For one GP a headline statistic on unplanned hospital admissions prompted reflection on his duty to prescribe safely:

“*…. when I read the blurb about this saying that 10% of hospital admissions could be caused by bad prescribing or ED attendances could be caused by bad prescribing. That’s pretty shocking and it’s quite an impetus to*-*you know*-*it really shows we have an onerous responsibility to be prescribing correctly.”* (GP5, male, partner-viewing high-risk NSAID prescribing)

### Core features and suggestions for improvement

Comparative benchmarking with peers or set standards was considered a core feature. The ability to compare results with those of their peers was motivating, although comparison with national standards was preferred. GPs were motivated by how their practice might be viewed by colleagues and did not want to be perceived unfavourably *(GP5, male, partner, Q9)*. One GP questioned whether being an outlier may in fact be a positive thing and may result in either complacency or disillusionment. However, GPs were also sceptical of feedback being used in a punitive way and preferred the idea of it supporting reflection and action rather than punishing poor performance *(GP2, female, partner, Q12)*.

The visuals of the dashboard were important with suggestions that graphs should be interpretable at a glance, with minimal text. It was possible to toggle between graphic and tabular display, and most GPs preferred graphic display. Some GPs found the lines in the trend graphs confusing: “*So first looking, this looks messy to me. It’s that is a bit overwhelming down in that part, but the bar charts I like.” (GP3, female, partner-viewing antibiotic prescribing)*. Several GPs had suggestions around adding a similar metric to the DU90% but instead of volume of prescribing to show cost *(GP2, female, partner, Q11)*. As well as highlighting cost, some GPs suggested breaking down the DU90% segment by therapeutic group or drug group and highlighting higher risk drugs within this.

Other GPs suggested additional information or resources that could add value, such as patient information leaflets on sick-day rules for high-risk medicines.

### Barriers

Despite feeling the dashboard was valuable and useful, all GPs reflected that with other pressures, they could not see themselves accessing it frequently. In fact, during the think-aloud process many GPs apologised that they had intended to view the dashboards in advance of the meeting but did not due to other demands.

### Engagement with the dashboard

GPs engaged with the dashboards in multiple ways, but predominately through an interpretative lens, particularly focusing on what the data meant in the context of their own practice. Interestingly many GPs responded with humour or anxiety, suggesting emotional stakes were high. Other GPs appeared less emotionally connected and commented more on design and layout and had suggestions for improvement. Some GPs showed scepticism, questioning data accuracy or methodological limitations (e.g., denominators, definitions). However, most took the data at face value, without questioning underlying assumptions even when limitations were footnoted (for example how the denominator was calculated and may be inaccurate). The three sub-themes that were applied deductively were interpretative, emotional and intentional engagement.

### Interpretative engagement

GPs generally engaged with the dashboards in an interpretative fashion. Some GPs initially struggled to interpret trend data but most quickly became used to the format after reading the axis labels and footnotes for the first graph, and became more heuristic in their interpretation, inferring meaning at a glance. Most GPs interpreted data in the context of their practice demographics *(GP6, female, partner-viewing practice DU90% where diazepam is the third most frequently prescribed drug, Q15)*, systems *(GP2, female, partner-viewing overall antibiotic prescribing rate, Q16)* and prescribers *(GP3, female, partner-viewing drugs in the practice DU90%, Q17)*. Many GPs commented on how hospital initiated prescribing influenced prescribing of specific drugs and how it was beyond their control. *“We just have to prescribe them because they’re prescribed in the hospital.*” *(GP5, male, partner-viewing red antibiotic prescribing)*. During think-aloud tasks, many GPs reflected on their own prescribing decisions, habits, or broader prescribing culture *(GP2, female, partner-viewing practice DU90%, Q18)*. Some GPs showed some scepticism, qualification, or challenge to the framing, assumptions, or usefulness of the dashboard data. Particularly with respect to high-risk NSAID prescribing; GPs were aware of the risk and felt there were other considerations not reflected in how the data was presented in the dashboard. *“You don’t know how long they’ve been given it for, like if somebody is on, you know, the ACE/ARB and diuretic and they get a very short course of ibuprofen, which they’ve tolerated before, maybe or maybe not.” (GP1, female, partner-viewing high-risk NSAID prescribing)*.

### Emotional engagement

Emotional responses ranged from relief to apprehension and disappointment to pleasure. Some GPs appeared surprised by what they saw, for example the volume of HRT prescribing: “*Estradiol. Yeah. More people are on HRT. God, we do a lot of it though, don’t we?” (GP1, female, GP-partner-viewing the practice DU90%)*. Negative emotions included resignation *(GP1, female, partner-viewing SSRI preferred drug prescribing, Q21)*, guilt *(GP5, male, partner-viewing high-risk NSAID prescribing, Q22),* disappointment (GP2, female, partner-viewing NSAID and anticoagulant prescribing*, Q23*) and anxiety *(GP6, female, partner-about to click on high-risk NSAID prescribing tab, Q24)*. The predominant positive emotion was relief. “*Yeah, the benzos [benzodiazepines] are pretty low. So that’s good.” (GP1, female, partner-BZRA prescribing rate)*.

### Intentional engagement

Intentional engagement was predominantly vague statements such as *“We probably need to tighten up a bit.”* (GP6, female, partner-viewing high-risk NSAID prescribing). Specific ideas included using the dashboard for practice quality improvement projects and as a starting point for discussions with trainees or colleagues. Intentional engagement tended to occur when GPs noticed they were an outlier or for higher risk metrics, or where GPs clearly indicated pre-existing interest in the metric.

### Safe, high-quality prescribing

GPs considered safe prescribing as protecting patients from medication-related harm *(GP8, female, assistant, Q26)*. High-quality prescribing was seen as balancing benefits and risks *(GP2, female, partner, Q27)*, with risks noted to be greater for older patients and marginalised groups. GPs recognised deprescribing as a necessary yet challenging process. GPs also considered financial implications as an important component of quality reflecting the “*finite pot or money available to the country” (GP2, female, partner)*.

In addition to hospital-initiated prescribing described above, GPs described other external factors that influence prescribing appropriateness such as patient expectations, pharmaceutical marketing, or inequity in access to care. Patients’ expectations regarding medications, was sometimes seen as a barrier to high-quality prescribing *(GP3, female, partner, Q6)*. With respect to commercial interests, one GP suggested that oral nutrition supplements (ONS) prescribing may be shaped by pharmaceutical company marketing, referring to a nurse promoting a specific product as *“Mrs Fortisip” (GP1, female, GP partner)*. The same GP suggested that ONS use in nursing homes may also be financially advantageous for the institution: *“If this is something that is prescribed, then they’re not paying for it, so it’s optimising a resident’s nutrition, but the nursing home doesn’t have to pay for it.” (GP1, female, GP partner)*.

## Discussion

### Summary of results

GPs were in favour of openness with prescribing data but were also worried about nefarious use by the pharmaceutical industry and by patients seeking out practices with higher opioid and benzodiazepine prescribing. GPs supported using prescribing data for feedback, with comparative benchmarking with peers or set standards identified as a core feature of this feedback. GPs emphasised that feedback should promote reflection and improvement, rather than feel punitive. Heavy workload was identified as a barrier, despite this, the dashboard was positively perceived and accepted by GPs. The predominant lens through which GPs engaged with the dashboards was contextual interpretation where they made sense of data based on their own staff, systems, prior quality improvement attempts and patient demographic. Many GPs showed a strong emotional attachment to their practice data and for some this was a motivator for reported intended behaviour change. Although all GPs reported an intention to act on what they saw, this was often vague. GPs considered safe, high-quality prescribing as enhancing effectiveness, minimising risk and being mindful of the financial burden their prescribing places on healthcare resources. Many GPs were interested in understanding how their prescribing compared in terms of costs, and this emerged as a frequent suggestion for enhancement.

### Comparison with the literature

Quasi-experimental and experimental studies have shown that simply accessing prescribing dashboards is associated with more cost-effective prescribing and that simple behavioural nudges can enhance the effectiveness of these dashboards (22, 23). Qualitative studies have shown how contextual factors such as trust and perceived agency influence engagement and how negative emotions such as apathy or resentment reduce the likelihood of behaviour change, particularly when feedback is perceived as irrelevant or as a form of external assessment (24, 25). Similarly, GPs in our study expressed a preference for feedback framed around reflection and improvement rather than punitive action. Apathy or resentment did not feature in our results. This may reflect social desirability bias but could also be because the interviewer, as a fellow GP, was perceived as contextually understanding and non-judgemental. Resignation or a sense that some issues could not be changed did emerge, particularly in relation to factors perceived as beyond a GP’s control such as hospital initiated prescribing or general overprescribing related to the perceived value of medicines in society at large.

However, a more striking feature was the emotional attachment many GPs demonstrated toward their prescribing data and a desire to improve and learn from their data.

Cost, and its relationship to high-quality prescribing, featured heavily in the results. GPs were interested in understanding how their prescribing contributes to the public purse and described this as a motivator for engagement. One policy response to this has been the HSE Preferred Drugs Initiative which identifies a single ‘preferred’ drug within a therapeutic class based clinical and cost-effectiveness (16). Prior research has shown poor adherence, despite the potential cost-saving benefits with improved uptake (26, 27). With shifts in prescribing patterns - including increased use of high-tech drugs and overall higher prescribing rates - policy attention is increasingly focused on these broader cost drivers (28). Nonetheless, GPs in this study were generally supportive of the Preferred Drugs Initiative. Many described making concerted efforts to switch to the preferred option once they were made aware of it. GPs also liked the idea of having a “go-to” drug within a therapeutic class aligning with principles outlined in the WHO Guide to Good Prescribing (29). With respect to drivers of non-cost-effective prescribing and overprescribing, GPs perceived undue pressure from the private sector and commercial interests as potential influencing factors. Funding models within general practice can also potentially influence prescribing but this did not emerge in this study. For example, capitation systems may increase the risk of sub-optimal or undertreatment, and fee per service funding models might incentivise overdiagnosis and overtreatment. Ireland has previous experience with financial incentives for “cost-effective” prescribing through the Indicative Drug Budget Scheme introduced in 1993, which returned a proportion of any savings to GPs’ practices (30). However the scheme was ultimately discontinued as although financial incentives shifted prescribing towards cheaper options this was sometimes with mixed implications for guideline-concordant care (30), suggesting cost should not be the sole organising principle when developing prescribing incentives or feedback systems.

### Strengths and limitations

The use of the think-aloud approach to capture real-time engagement is a major strength of this work. Similarly, triangulation of the think-aloud and semi-structured interviews provided both immediate reactions and more reactive insights. The interviewer (CMC) is a practicing GP; this may have enhanced the depth and nuance of the data collected and interpreted. At the same time, the involvement of a non-clinical co-investigator (AE) brought a valuable external perspective.

This study has some limitations, this was a small relatively select sample of GPs in a single healthcare context, over-represented by practice partners. However, practice partners may have more ability to influence prescribing at the practice level, so capturing their views is paramount. Recruitment was slow and did not reach the number outlined in protocol, with time constraint the most commonly cited reason for non-participation.

### Implications for practice, policy and research

Addressing overprescribing and safe prescribing is a key policy initiative in many health systems worldwide (4, 5). Unwarranted variation in prescribing - that is, variation not explained by patient need- is a marker of suboptimal care (31). Increasingly, this variation can be identified in near real-time through electronic health records and national prescribing databases, offering an opportunity for quality improvement (32). This study describes how GPs both engaged with and perceived this feedback. Overall, any form of comparative feedback was welcomed; GPs actively encouraged feedback and were very reflective in their response to it. A pragmatic approach would be to work iteratively in the constraint of what is possible within current systems to facilitate this for clinicians. A major barrier in the Irish context is the two-tiered system where private prescribing is not routinely captured in any database. In addition, data governance issues around access to data need to be addressed. Recent research in this area has highlighted the complex and often contradictory views of various stakeholders (33). However, as one participant aptly described, that although our caution is due to concerns over data privacy, lack of transparency does nothing to protect the patient. *OpenPrescribing.net* is a prominent example of how open practice level prescribing data can be used for comparative feedback (11). More granular metrics that require patient-level data need more complex analytic approaches such as the federated approaches used by *OpenSAFELY* (34).

## Conclusions

GPs were open to using prescribing dashboards when the information was clear, relevant and supportive rather than punitive. Prioritising a small number of high-volume, high-cost and high-risk indicators may offer a practical starting point. System-wide, such an approach has the potential to improve patient outcomes and reduce costs. However, this potential will only be realised if dashboards are co-designed from the outset to maximise engagement. In addition, the impact of any system-wide prescribing dashboards should be formally evaluated. In the mean-time identifying opportunities for improvement within current systems is important. This study provides insights to inform future dashboard development, while also showing how a locally driven, innovative approach can directly support safe and effective prescribing.

### Patient, public and knowledge user perspectives

In March 2023, at the grant proposal stage, the PI (CMC) met with two public and patient involvement (PPI) contributors to discuss the overall research aims of the proposal and how they would be addressed. Their feedback helped shape the initial proposal. The grant’s independent steering committee includes a GP representative, and the PI is also a practicing GP, ensuring that clinical perspectives are embedded throughout the project.

## Data Availability Statement

Transcripts are not available as participants were not asked for consent for sharing them.

## Extended data

The study protocol is available at https://hrbopenresearch.org/articles/8-67/v1. The think aloud tasks, interview guide and SQL code used to generate variables for visualisation in QuickSight are available at: https://doi.org/10.5281/zenodo.15545093

## Ethical approval and consent to participante

Ethical approval was granted by the Irish College of General Practitioner Research Ethics Committee, 7^th^ October 2024, ICGP_REC_2024_ 2502. All participants gave fully informed written consent.

## Consent for publication

Not applicable.

## Author contributions

All authors made substantial contributions to the work, including drafting, writing, or critically reviewing the manuscript, and take responsibility for the content.

AS analysed and interpreted the semi-structured interview data and prepared the first draft of the manuscript. BC interpreted the analysed data, provided methodological guidance, and critically reviewed the manuscript. TF provided methodological guidance and critically reviewed the manuscript. FM interpreted the analysed data, provided methodological guidance, and critically reviewed the manuscript. MF observed the pilot think-aloud and interview process, contributed to refinement of the interview schedule, interpreted the analysed data, provided methodological guidance, and critically reviewed the manuscript. CG interpreted the analysed data, provided methodological guidance, and critically reviewed the manuscript. CMC conceptualised the study and methods, conducted the think-aloud and semi-structured interviews, analysed the think-aloud data, interpreted the data, prepared the first draft, and critically reviewed the manuscript.

All authors read and approved the final manuscript.

